# Towards a Social Media-based Disease Surveillance System for Early Detection of Influenza-like Illnesses: A Twitter Case Study inWales

**DOI:** 10.1101/2024.11.11.24316812

**Authors:** Mark Drakesmith, Dimosthenis Antypas, Clare Brown, Jose Camacho-Collados, Jiao Song

## Abstract

Social media offers the potential to provide detection of outbreaks or public health incidents faster than traditional reporting mechanisms. In this paper, we developed and tested a pipeline to produce alerts of influenza-like illness (ILI) using Twitter data. Data was collected from the Twitter API, querying keywords referring to ILI symptoms and geolocated to Wales. Tweets that described first-hand descriptions of symptoms (as opposed to non-personal descriptions) were classified using transformer-based language models specialised on social media (BERTweet and TimeLMs), which were trained on a manually labelled dataset matching the above criteria. After gathering this data, weekly tweet counts were applied to the regression-based Noufaily algorithm to identify exceedances throughout 2022. The algorithm was also applied to counts of ILI-related GP consultations for comparison. Exceedance detection applied to the classified tweet counts produced alerts starting four weeks earlier than by using GP consultation data. These results demonstrate the potential to facilitate advanced preparedness for unexpected increases in healthcare burdens.

## 1 Introduction

Surveillance of symptoms of infectious diseases in the population, also known as syndromic surveillance, is an important public health function to provide warning of an incoming epidemic and prepare healthcare systems for increased demand. Such surveillance systems traditionally rely on clinical data, for example GP consultations, ambulance call-outs, sickness-related absences, and access to telephone advice services. Such data sources can be slow due to the reporting mechanisms they rely on. In recent years, there has been increasing interest in leveraging social media to develop an early warning detection (EWD) system for infectious diseases (Rodríguez-Martínez and Garzón-Alfonso, 2018; Espinosa et al., 2022; Şerban et al., 2019; Aiello et al., 2019; Joshi et al., 2019; Ofoghi et al., 2015). Social media provides a rapid, and high-volume data source, offering the potential to provide reliable detection of a disease outbreak or public health incident faster than traditional reporting mechanisms. Such data sources will enable more rapid and timely response to anticipate increased demand on health services.

We developed a pipeline to ingest Twitter (now known as X) data matching symptom-related keywords, localised to Wales, classify tweets that describe first-hand accounts of influenza-like illnesses (ILIs), and apply an exceedance detection algorithm to produce alerts of higher-than-expected incidence. We test the method’s ability to provide early warning of the recent spike in flu cases in the end of 2022 which was much higher than in the previous years and placed significant demand on the healthcare system. We compared the performance of the method applied to Twitter data to that applied to GP consultations for ILIs, a more established syndromic surveillance indicator.

## 2 Methods

In order to detect potential outbreaks in social media, we primarily rely on Twitter data (Section 2.1) and an automatic NLP-based classification methodology (Section 2.2). Then, we present the GP consultation data we use as a comparison (Section 2.3) and our methodology to identify outbreaks based on data (Section 2.4).

### 2.1 Data

The Twitter Academic API (Twitter, 2023) was utilised to collect tweets originated from Wales from January 2020 to January 2023. Only tweets in the English and Welsh language were collected.

#### 2.1.1 Geolocation

Due to limitations of Twitter’s API (i.e. we could only filter countries by ISO alpha-2 code and there is no one available for Wales) we utilise a map of Wales (https://datashare.ed.ac.uk/handle/10283/2410?show=full) and divide it in 34 equal areas which are used as parameters for the “boundary_box” field of the API. In total, 5,278,425 tweets were gathered that certainly originated from the region of Wales.

#### 2.1.2 Keyword matching

In an effort to collect a sufficiently large dataset that can allow us to identify any early signal related to flu outbreaks, we initially identified relevant tweets by applying a filter of 22 relevant keywords during the API call: (’flu’, ’ill’, ’sick’, ’unwell’, ’fever’, ’cough’, ’coughing’, ’bug’, ’headache’, ’hoarseness’, ’muscle pain’, ’sore throat’,’high temperature’, ’tummy pain’, ’covid-19’, ’covid’, ’covid19’, ’coronavirus’, ’blocked nose’, ’runny rose’, ’aches’, ’fevery’). Keywords referring to symptoms were principally used, but also keywords referring to ’flu’ and ’covid’ were included as these are likely to be mentioned in lieu of respiratory symptoms. With geolocation and keyword filtering, 35,724 tweets were retrieved. Counts and overview of the size of the datasets can be found in Figure 1 and Table 1.

**Table 1:** Counts of tweets pulled pulled from the API, with geolocation, keyword matched and classified for first-hand accounts of ILI symptoms. (2022 includes 1st week of 2023)

| Year | Geolocated to Wales | Matching ILI keywords, without classification | Classified as first-hand account of ILI symptoms |
| --- | --- | --- | --- |
| 2020 | 1,382,874 | 13,173 | 499 |
| 2021 | 1,967,665 | 14,225 | 823 |
| 2022 | 1,927,886 | 8,326 | 891 |
| Total | 5,278,425 | 35,724 | 2,213 |

**Figure 1:**
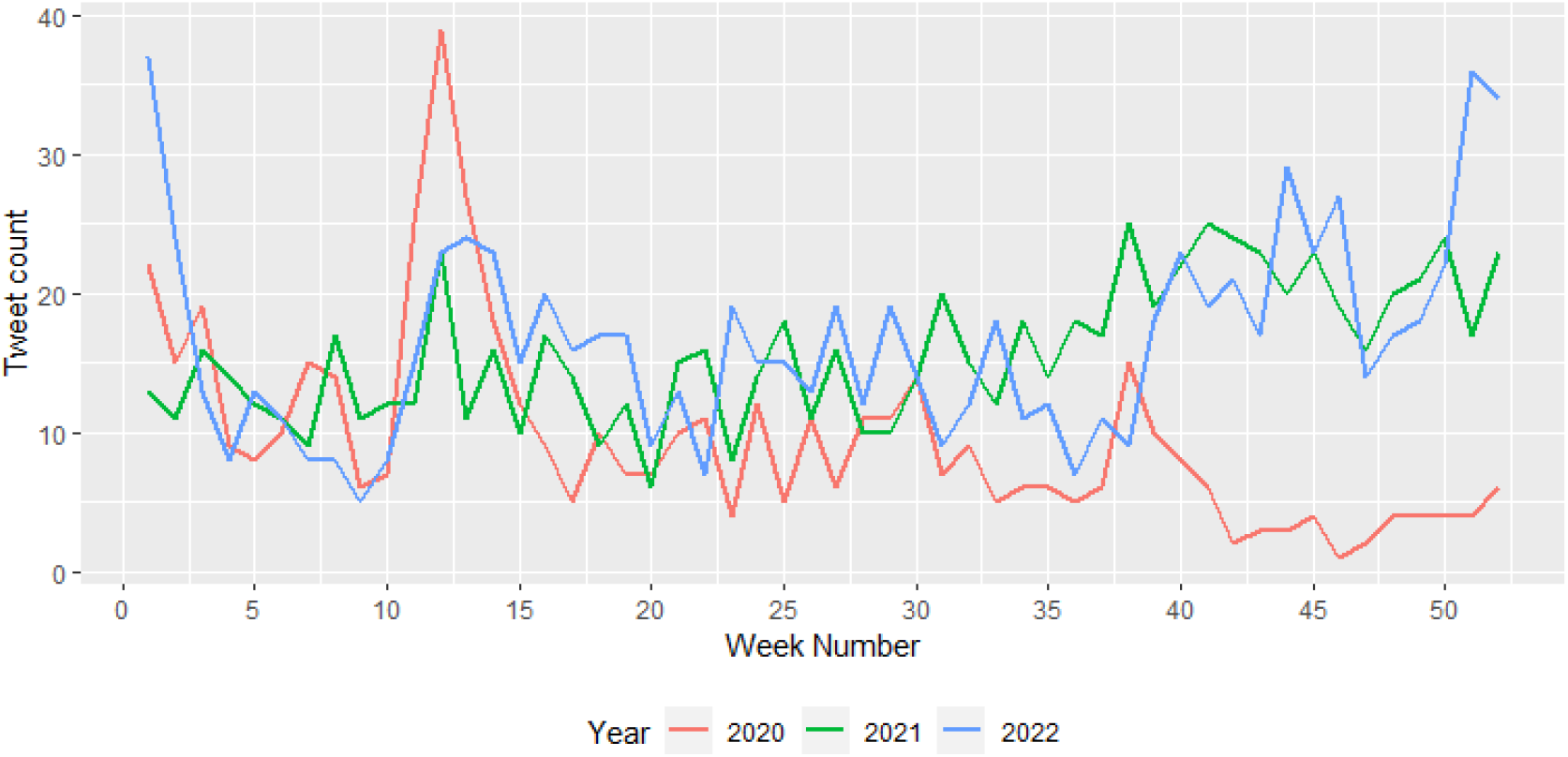
Counts of tweets classified as referring to first-hand accounts of ILI symptoms, geolocated to Wales.

### 2.2 Classification

An important consideration is the identification of first-hand accounts of symptoms being experienced, which are more representative of prevalence of symptoms in the community, as opposed to tweets that provide general advice or discussion about symptoms and illnesses (see Table 2 for examples). We therefore employ a small manual annotation (section 2.2.1) and NLP methods (Section 2.2.2) to classify such tweets. In total, 2,213 tweets were classified as being first-hand accounts, approximately 6% of all retrieved tweets.

**Table 2:** Examples of tweets classified at whether or not they describe first-hand accounts of ILI symptoms.

| Class | Example |
| --- | --- |
| TRUE | "I've never felt so ill. coughing, shivering, aching. Luckily my mom is supplying me with medication, water and soup!" |
| FALSE | "If you have aches, fever and feel generally unwell, it might be flu. Make sure to rest, drinking plenty of water & take over the counter medicines to ease symptoms." |

#### 2.2.1 Manual Annotation

Aiming to investigate the difficulty of the classification and also to collect data for the training of machine learning models a sample of 121 tweets was also manually annotated by three different coders. This sample was geolocated to the whole UK, to avoid a large overlap with the Wales dataset to serve as an independent training set. Each annotator was asked to annotate tweets as TRUE if the tweet contained descriptions of ILI symptoms from a first-person perspective, and FALSE otherwise. Our results indicate a strong agreement between the coders which achieve an average of 0.68 when considering Cohen’s Kappa. Having established a high agreement between the coders, an additional 751 tweets were individually annotated, bringing the total number of gathered tweets to 878^1^.

#### 2.2.2 Automatic Classification

There has been a recent influx of new large language models in the NLP field such as OpenAI’s GPT-3 and Google’s Bard that achieve impressive results on difficult tasks. However, for simpler tasks such as binary text-classification, as in our use case, smaller models fine-tuned for the particular task can achieve similar performances without the need of huge computational resources or paying API services.

Two large pre-trained language models were tested for the classification process: Twitter-RoBERTa (Loureiro et al., 2022) and BERTweet (Nguyen et al., 2020). Both models are based on the RoBERTa (Liu et al., 2019) architecture which in turn is an expansion of BERT (Devlin et al., 2019). RoBERTa models are essentially deep neural networks of 12 layers and utilise techniques such as attention masks (Vaswani et al., 2017) and dynamic masking of tokens during training that allows them to understand language relations and create accurate representations (by mapping words into a high dimension embedding vectors). To achieve this the models are usually trained on large corpora of text. Specifically, BERTweet and Twitter-RoBERTa are pre-trained in a large corpus of tweets, 850 and 124 million tweets respectively, and are tailored for usage in social media context.

These transformer-based language models were selected as: (1) they consistently outperform previous text-classification approaches and adapt well to different domains and (2) since they have been pre-trained on Twitter data, they perform better in social media text data (i.e. short, unstructured text, internet slang, emojis, etc) (Barbieri et al., 2020; Antypas et al., 2023).

Due to the small size of the annotated set, a 5-fold cross-validation method is also applied where the whole dataset is used. We also ensured that distribution of classes in the train and test sets in each fold is the same.

All models used are based on the implementations of the base versions provided by Hugging Face (Wolf et al., 2020) while Ray-Tune (Liaw et al., 2018) was utilised to optimise the models’ hyper-parameters (i.e. learning rate, training batch-size, warm-up rate, number of epochs).

To establish the difficulty of the task and better evaluate the performance of the language models, three baseline models were also tested. An SVM classifier based on TFIDF features is tested, along with two frequency-based classifiers (predicting the most frequent and least frequent classes).

### 2.3 Clinical data

As a comparison to the Twitter data, a more traditional public health indicator was also analysed. Data on weekly counts of GP consultations for influenza-like illnesses (ILI) reported in Wales for the period January 2020 to December 2023 was extracted (Public Health Wales, 2023). A total of 11,152,985 GP consultations for ILIs were recorded for the period of interest. There was a notable surge in ILI-related consultations in towards the end of 2022, peaking in weeks 49-51 (Figure 2)

**Figure 2:**
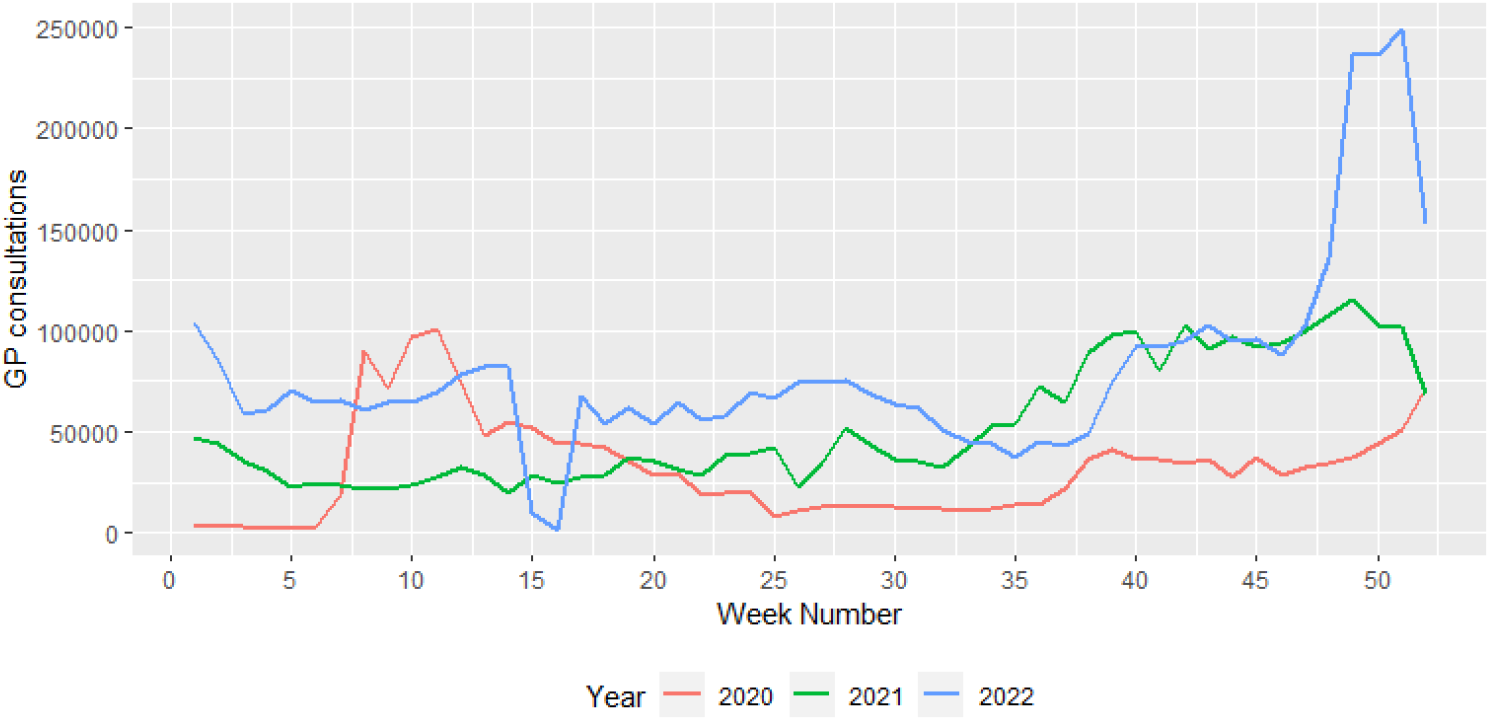
Counts of ILI-related GP consultations between 2020 and 2022

### 2.4 Exceedance detection

Exceedance detection was performed using a modified version of the Farrington algorithm (Noufaily et al., 2013) as implemented in the R package *surveillance* (Salmon et al., 2016). Briefly, the algorithm iteratively fits a quasi-Poisson model to historic data and detect significant deviations in the present data from that predicted by the model. The model was fit to the 2 previous years of data (2020-2021) and exceedance detected for data in the year 2022. Counts were binned into weekly intervals. A window width or 3 weeks and detection threshold of *α* = 0.05 (all other parameters kept to default values).

Exceedance detection was applied to the classified tweet counts. For comparison, the keyword-matched tweet counts (without classification) and the counts of ILI GP consultations were also analysed.

## 3 Results

In this section, we present both the classification results that are used as basis for our analysis (Section 3.1), and the results of our exceedance detection algorithm based on the classification output (Section 3.2).

### 3.1 Classification results

When considering the results of the cross-validation classification experiment, the BERTweet model performs slightly better than the Twitter-RoBERTa (T-RoBERTa) one. Table 3 displays the average macro-F1 scores for each class that the two models achieve.

**Table 3:** Average F1 scores for each class. The accuracy and mean macro-F1 scores are also reported. The “Most Frequent” baseline indicates a baseline predicting always FALSE, while the “Least Frequent” baseline indicates a baseline predicting always TRUE.

|  | F1 |  |  | Accuracy |
| --- | --- | --- | --- | --- |
|  | FALSE | TRUE | Macro |  |
| **BERTweet** | **88.44** | 74.70 | **81.57** | **84.16** |
| **T-RoBERTa** | 87.30 | **74.80** | 81.05 | 83.14 |
| **SVM** | 84.13 | 33.39 | 58.76 | 74.37 |
| **Most Frequent** | 82.23 | 0.00 | 41.11 | 69.82 |
| **Least Frequent** | 0 | 46.37 | 23.18 | 30.18 |

Both models appear to struggle to identify the positive class where their performance drops approximately by 10 ten points in terms of F1 score. In general, tweets that indicate first-person flu symptoms in a more subtle way pose difficulties for the models. For example both *‘When you’re too ill to watch TV or read a book music is what keeps you sane. Currently listening to Queen. Freddie was the absolute best, without a doubt*.*’* and *‘@user My family had it we were ill for 2 days 7 of us non jabbed*… *‘* are labelled as positive but classified by BERTweet as negative entries. At the same time, negative labelled tweets such as *‘@user My partner has had it for 2 feverish nights horrendous coughing, headache, sore eyes but covid negative*.*’* and *‘Is there anyone in this country NOT coughing and spluttering and snotting and just generally feeling yuck*¿ which describe symptoms in a more general way are also labelled as positive by BERTweet.

Due to its slightly better performance, the BERTweet classifier was used for subsequent analysis.

### 3.2 Exceedance detection

Using the classified tweet counts provided by BERTweet, a cluster of alerts was triggered for weeks 44-46 and 50-52 of 2022 (Figure 3 – see Section 2.4 for more details on how this experiment was performed). For ILI GP consultations, a cluster of alarms were triggered for weeks 48 of 2022, onwards (Figure 5). The classified tweet counts therefore alerted to a significant increase in ILI-related tweets approximately 4 weeks before a corresponding alert was triggered for ILI-related GP consultations. For tweet counts derived from ILI keyword matching without classification, no alerts were triggered (Figure 4).

**Figure 3:**
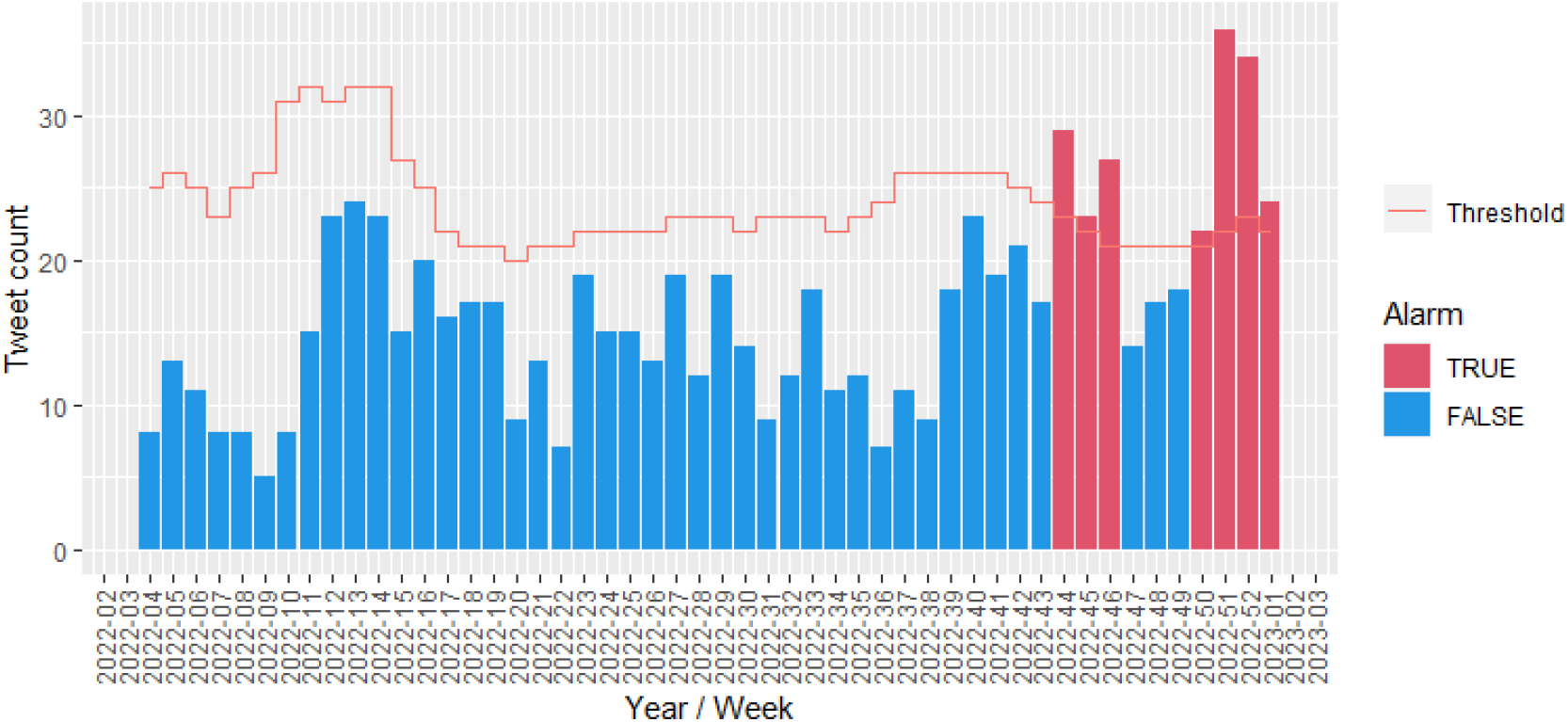
Exceedance detection of weekly tweet counts in 2022, geolocated to Wales, classified as mentioning first-hand accounts of ILI symptoms

**Figure 4:**
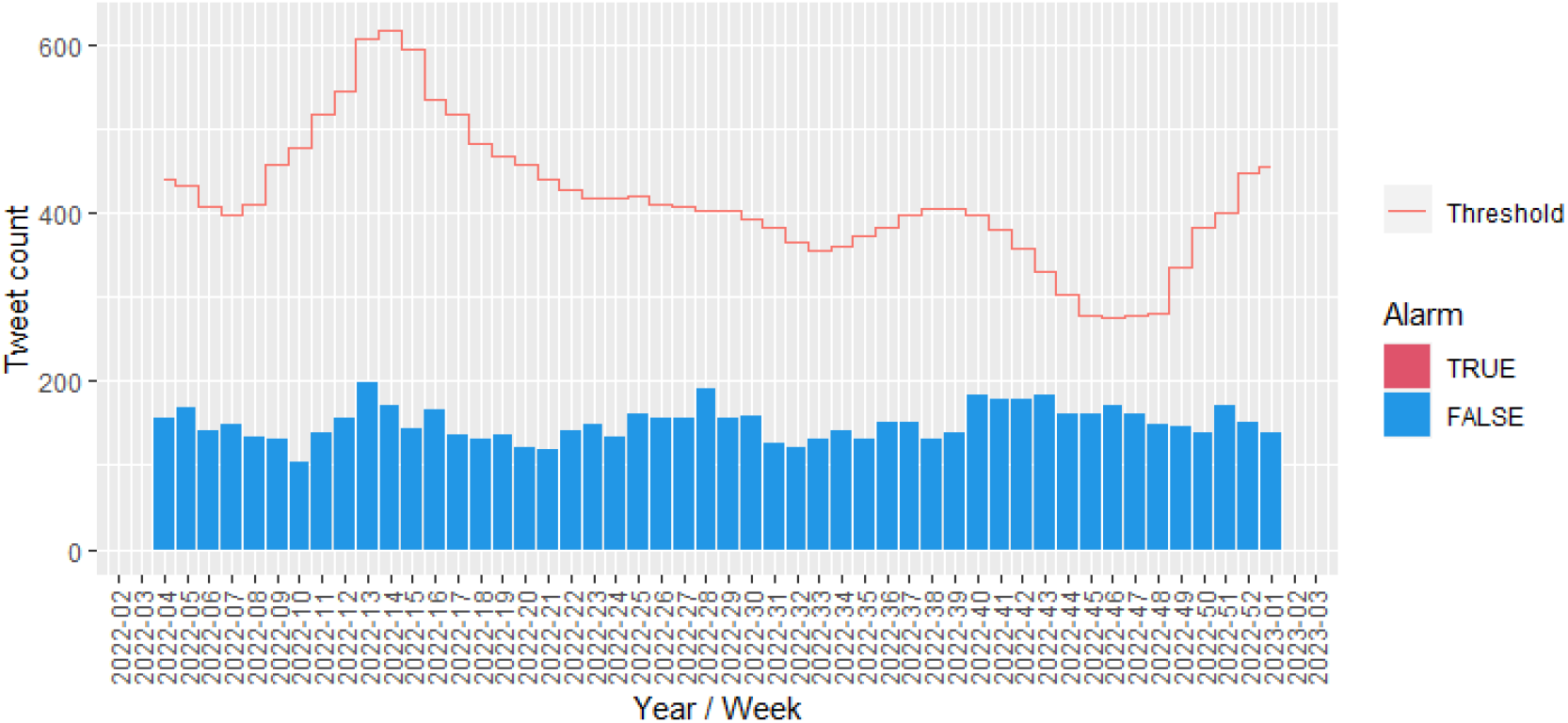
Exceedance detection of weekly tweet counts in 2022, geolocated to Wales, matching ILI keywords, without classification.

**Figure 5:**
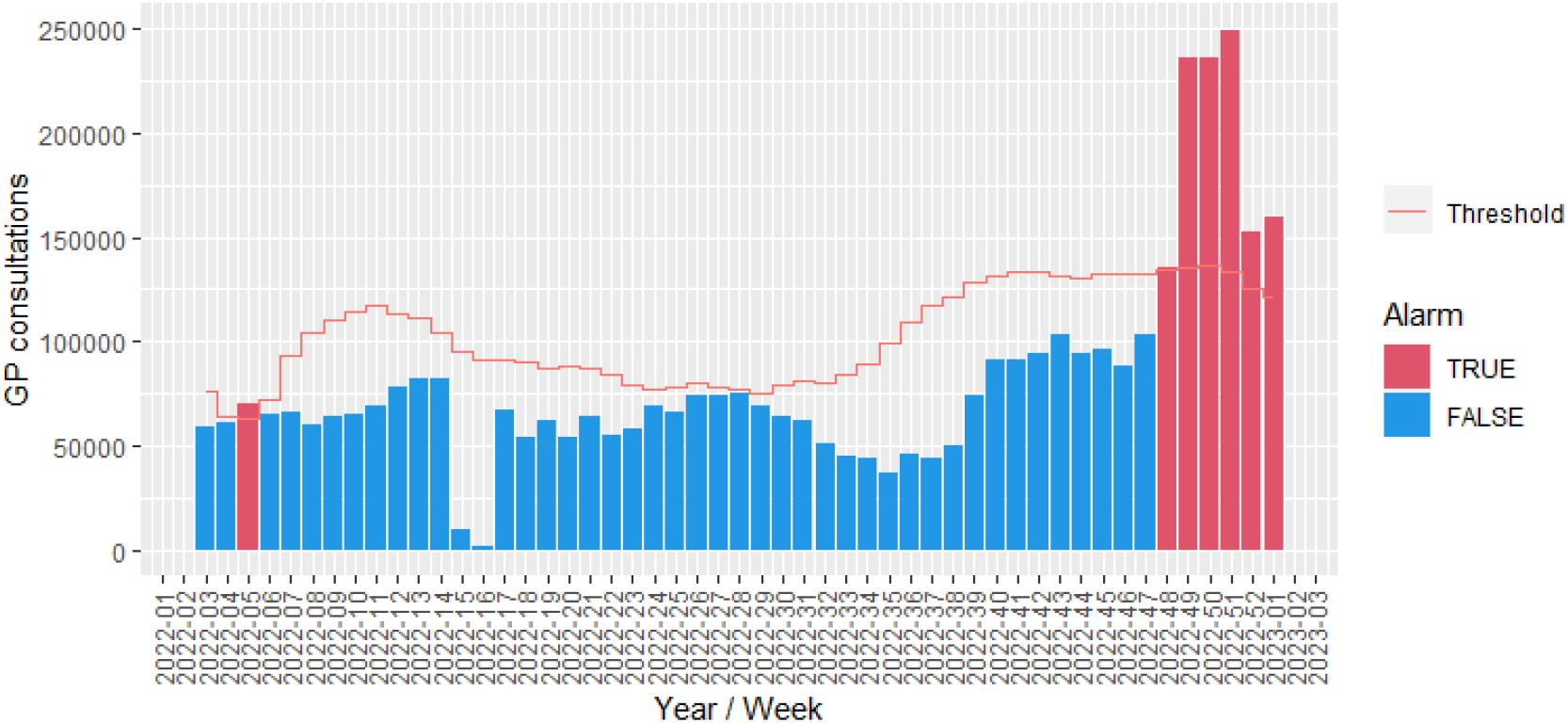
Exceedance detection of ILI-related GP consultations in 2022 in Wales.

## 4 Discussion

This study has demonstrated the utility of using social-media data to provide early exceedance alerts of influenza-like illnesses (ILIs) earlier than routine clinical data. the classified tweet counts produced an exceedance alert 4 weeks lead time on routine clinical data. It can supplement existing evidence for practitioners to assess if a season has started earlier than anticipated or is more extreme than usual, and provide public health authorities a valuable tool to prepare for an incoming surge in demands on the healthcare system.

Furthermore, this study shows the importance of correctly classifying first-hand accounts of symptoms, as matching on ILI-related keywords alone produced counts that are heavily biased by mass media content, not reflective of prevalence of symptoms in the community. We show that tweet counts using keyword matching alone failed to produce any alarms. Despite the fact that the classifier struggles with some particular linguistic patterns and contexts, the classified tweet counts does show a significant increase in tweet counts that coincide with an increase in GP consultations.

### 4.1 Limitations

A significant issue is the relative scarcity of Twitter users who enable geolocation. Approximately 3% of tweets matching keywords had geolocation enabled. An attempt was made to overcome this retrieving data without geolocation and estimating location with the *carmen* Python library. This method had its own issues due to the large volume of key-word matching tweets exceeding the available quote of the API, but then very few located to Wales, much fewer than using true geolocation. The resulting performance was poorer.

The exceedance detection methods has limitations. It is only useful for detection of exceedance in infections with seasonal trends (e.g. influenza). It can only detect increases higher than on previous years, but does not provide a quantification of the magnitude of the increase. An alternative approach is to use time-series modelling, such as ARIMA, GAM, etc. which do not rely on seasonal behaviour. A further issue is that the approach to EWD, is that only significant increases, relative to previous years will be detected. An increase in healthcare burdens may be operationally significant within a given year, even if it is not statistically higher than previous years. The approach however, is useful for detecting increases in healthcare burdens that are unusually early compared to previous years.

The choice of training dataset has potential issues. There may be language differences between Wales and the rest of the UK, either use of Welsh language or Welsh-specific dialects in English specific to Wales, which the training dataset would not capture.

For robust exceedance detection, it is recommended to use 5 years of data to compute the baseline (Noufaily et al., 2013). However due to the API quota limits on how many historic years’ data could be retrieved, we could only use 2 years (2020-2021). This makes it difficult to comprehensively evaluate the reliability of the method. A further complication is that these years were significantly impacted by the Covid-19 pandemic. This is likely to lead to significant changes in how people engage with social media compared to normal.

While the proposed approach shows promise for providing EWD for infectious diseases, a significant setback occurred with the withdrawal of the free Academic Twitter API in early 2023. This makes this data source much less accessible. This has big implications for research and non-commercial applications such as disease surveillance (Davidson et al., 2023a,b). There are a number of alternative ’microblogging’ platforms that have received more attention in the past year, such as Mastodon, Threads and Bluesky. However, these platforms do not have the same volume of users as Twitter, meaning content relevant for syndromic surveillance will be very sparse. Also, most of these platforms lack an accessible interface to facilitate rapid automated data retrieval. Platforms such as Instagram and TikTok have very large user-bases, but the predominance of image and video-based content makes them unamenable to NLP methods. More generally, changing patterns in how the public engage with social media should be considered. In recent years concerns about privacy and digital sanctity have driven social media users to be less inclined to publicly share details of their personal well-being. A 2018 survey of social media use in Wales reported only approximately 10% of users shared details of their health on social media, and only a quarter of those were shared publicly (Song et al., 2020). It is expected that this proportion will be lower today. Another study monitored Twitter usage among participants in a flu symptom survey (Daughton et al., 2018). This study found participants rarely tweeted about their symptoms while experiencing them (of 266 symptom-related tweets identified, only 3 were made within 2 weeks of an instance of flu-like symptoms, of which there were 426). If these usage patterns are reflective of the wider population, it will significantly impact on the reliability of social-media as a syndromic surveillance tool.

Digital representativeness is another issue. Social media platforms disproportionately over-represent some demographics over others (Anderson, 2021). In Wales, there was no significant difference in social media use across demographics, although, use of Twitter was lower in more deprived areas (Song et al., 2020). The potential for social media to reach hard-to-reach populations should be considered for distributing important public heath guidance to control the spread of disease.

## 5 Conclusion

In this paper, we demonstrate that social-media based syndromic surveillance is capable of providing an advance warning of healthcare burdens earlier than traditional syndromic indicators. The results are encouraging and suggest that an adoption of social media indicators to supplement traditional disease surveillance is feasible with current technology. We also demonstrate the importance of NLP-based classification in identifying references to first-hand accounts of symptoms experienced. Nonetheless, validation of these conclusions with larger datasets is warranted. Furthermore, issues around access to social media data, digital representation and changing patterns of social media engagement should be considered when using social media data for syndromic surveillance.

## Data Availability

All data produced in the present study are available upon reasonable request to the authors

## 6 Ethics Statement

Twitter data was accessed via the Twitter Academic API (Twitter, 2023) and only aggregated anonymised data is presented. Additionally, all user mentions have been removed from posts. No individual health status data was reported or analysed.

Aggregated GP consultation data is openly available from the Public Health Wales ARI GP Consultations dashboard (Public Health Wales, 2023).

## Footnotes

1 Class distribution: 613 FALSE, 265 POSITIVE entries.

